# Clinicopathological features and outcome of COVID-19- early experiences from three covid hospitals, Chittagong, Bangladesh

**DOI:** 10.1101/2021.05.11.21256930

**Authors:** Rajat Sanker Roy Biswas, Jishu Deb Nath, Pranab Kumar Barua, Md Rejaul Karim, Safatujjahan, Mohammad Saiful Islam, Kazi Farhad Ahmed

## Abstract

**Introduction:** COVID 19 is an unknown virus affecting mankind creating a deadly experience to all. It is true for Bangladesh also. So the objectives of the present study is to find the clinicopathological features and outcome of COVID patients admitted in three COVID dedicated hospitals of Chittagong, Bnagladesh.

**Methods:** This was an observational study where a total of 209 patients admitted in three COVID dedicated hospital were recruited. Clinicopathological data were recorded and patients were under observation till discharge and thus outcome were recorded. Prior consent was taken from the patients and ethical clearance was also taken. Data was compiled and analyzed by SPSS-20.

**Results:** Among 209 patients most of them were male 139(66.5%) and male to female ratio was 1.98:1. Age group distribution revealed more were aggregated in age group 41-50 years 36(17.2%), 51-60 years 54(25.8%) and 61-70 years 57(27.3%). Among all 92(44%) patients were RT-PCR positive and 117(56%) were probable cases. Fever was present in 195(93.3%) cases, cough in 180(86.1%), respiratory distress in 105(50.2%) anosmia in 123(58.8%), aguesea in 112 (53.58%) and lethargy was present in 143(68.42%). Chest X-ray findings revealed 73(34.9%) had bilateral patchy opacities, 20(9.6%) had unilateral opacities 65(31.1%) had consolidations, 6(2.9%) had ground glass opacities and 2(1.0%) had pleural effusion. Supplemental O2 was given in 173(82.8%) patients, Favipiravir in 59(28.2%), Remdisivir in 111(53.1%), Methylprednisolone in 87(41.6%), Dexamethasone in 93(44.5%), Antibiotics in 204(97.60%), Toccilizumab in 34(16.3%), plasma in 18(8.6%) and LMWH in 200(95.7%) patients. Regarding outcome of the COVID patients admitted, 85(92.4%) patients improved, 6(6.5%) died who were RT-PCR positive and 107(91.15%) improved, 9(7.7%) died who were probable cases. Total death rate was 7.1%.

**Conclusion:** Present study findings were some early activities among COVID patients in the years 2020. Male were more affected and middle age group people were the most victims.

## Introduction

In December 2019 a new respiratory tract infecting agent emerged in Wuhan city of China, known as the coronavirus.^1^ It was later named Covid-19. COVID-19 has now become a pandemic. While the origin of the 2019-nCoV is still being investigated, current evidence suggests spread to humans occurred via transmission from wild animals illegally sold in the Huanan Seafood Wholesale Market.^1^

Virus spread rapidly through China infecting more than 85,000 people. Within a few months it engulfed the Europe causing massive loss of life and property in Italy, Spain, France, Germany, UK and then USA. It is now spreading in Bangladesh which is one of the populous country of the world in relation with total land areas.^2^ As of now more than 600900 people have been infected and 8950 people have succumbed to the illness in our country till March 2021 in Bangladesh.^3^

The WHO declared Covid-19 a global pandemic on 11March 2020. Illness ranges in severity from asymptomatic or mild to severe; a significant proportion of patients with clinically evident infection develop severe disease. Human-to-human transmission via droplets as well as through contact with fomites act as routes of the virus spread. Among the infected populations 80% are either asymptomatic or have mild disease, people have been going to their workplaces and even traveling internationally. Nevertheless, even though the virus is causing mild disease in many, the course of illness may be severe, leading to hospitalization and even death in elderly or those with comorbid conditions.^4^

Guan et al.^4^ published a report on 1099 patients with laboratory confirmed Covid-19 from 552 hospitals in China through January 29, 2020. The most common symptoms reported were fever (43.8% on admission, and 88.7% during hospitalization) and cough (67.8%), diarrhoea (3.8%) was uncommon. A severe form of the disease was reported in elderly and in patients with comorbidities. Mortality rate among diagnosed cases (case fatality rate) has a variable range; true overall mortality rate is uncertain, as the total number of cases (including undiagnosed persons with milder illness) is unknown

Data of covid is under way from the different parts of world but it is still scarce from Bangladesh So objectives of this paper is to describe the clinical profiles of covid patients ranging from their age, sex, clinical symptoms, laboratory evaluation, radiological characteristics and treatment provided along with outcome.

## Methods

In this observational study, we included 209 cases of reverse transcription polymerase chain reaction (RT-PCR) positive COVID-19 patients as confirmed cases and those with RT PCR negative but with supportive clinical history and radiological evidences as probable cases. Data were collected between June to December, 2020 from three COVID dedicated hospitals Chittagong Bangladesh. A structured questionnaire was used to collect the data and all patients were observed till discharged irrespective of outcome. Eventually 209 cases were enrolled. Informed written consent was obtained from every patient or from legal guardian by reading out according the revised Declaration of Helsinki. The protocol was approved by the Ethical and Scientific Committee of the Chattogram Maa O Shishu Hospital Medical College (CMOSHMC). We collected demographic data (age, sex, etc.), clinical data (symptoms on admission, investigations reports etc.) and correlated them with outcome. The statistical analysis was carried out using the Statistical Package for Social Sciences version 20.0 for Windows (IBM SPSS Armonk, NY, USA). Qualitative variables such as fever, cough etc. were expressed as frequency and percentage. Quantitative variables were expressed as mean ± standard deviation.

## Discussion

First COVID-19 cases were declared by Bangladesh in Dhaka City on 8 March, 2020, highest number of cases have been detected in Dhaka3 and thus it is considered as the core of the disease transmission in Bangladesh. Since then covid cases increased gradually.^5^

Among 209 patients studied most of the patients were male139(66.5%) and male to female ratio was 1.98:1. It was similar to that reported by Huang et al^6^ and Chen et al^1^ which show 73.0% male predominance but higher than that reported by Wang et al^7^ (54.3%). This male predominance may have happened due to increased foreign travel by males fo occupational or educational purposes.

Age group distribution revealed more were aggregated in age group 41-50 years 36(17.2%), 51-60 years were 54(25.8%) and 61-70 years 57(27.3%). Our socio-demographic findings, matched that of Asia, e.g. China^8^ (median age: 47 years; 41.9% female), India^9^ (mean age 40.3 years, 66.7% male) and other reports from Bangladesh^10^ (43% were in the age range of 21 to 40 years, female: male ratio 1:2.33). But studies from America8 (median age, 63 years) and Europe9 (Median age, 67.5 years) showed higher age of patients but same male preponderance.

Among all 92(44%) patients were RT-PCR positive and 117(56%) were probable. As per case definition RT-PCR positive cases were taken as confirmed cases and who had clinical and radiological findings compatible with COVID 19 were taken as probable cases. RT-PCR was the first line of diagnosis in patients with COVID-19 in Bangladesh. In previous reports, chest CT scan was found to be a more sensitive diagnostic tool than RT-PCR even in asymptomatic patients reaching 98%.^12^ However, many researchers found that patients with a positive RT-PCR may have a negative chest CT scan, and patients with a negative RT-PCR may have positive chest CT scan.^12^ Chest x-ray was regarded an insensitive tool reaching 69%.^12^

Clinical findings revealed fever was present in 195(93.3%) cases, cough in 180(86.1%), respiratory distress in 105(50.2%) anosmia in 123(58.8%), aguesea in 112 (53.58%) and lethargy was present in 143(68.42%). In our study fever and cough was the most common symptom present in our patients which was similar to that reported in Huang et al^6^ and Wang et al^7^ where fever and cough also were two common symptoms found. Some patients were found asymptomatic also.

The some most common chest x-ray finding in our patients were bilateral and unilateral patchy opacities and GGO in a peripheral distribution, there was a lower lobe predilection of the opacities, with the right lower lobe more common than the left lower lobe. Our findings are in consensus with previous studies on chest x-ray and chest CT scans. ^11,12^ Only two patients had pleural effusion which is not a common finding on chest imaging^13^ in our study the presence of symptoms correlated significantly with abnormal chest x-ray findings suggesting that chest x-ray may be helpful as an aiding tool in the diagnosis and follow up in patients with COVID-19 pneumonia.

Laboratory parameters were variable among confirmed cases and probable cases. CRP, Ferritin and d-dimer were used to check as inflammatory markers and hematological parameters were also reviewed. Furthermore, blood hypercoagulability is common among hospitalized COVID-19 patients. Elevated D-dimer levels were consistently reported as well. Thus, the study concluded that in patients with COVID-19 either hospitalized they are at high risk for venous thromboembolism, and an early and prolonged pharmacological thromboprophylaxis with LMWH is highly recommended.^14^

Supplemental O2 was given in 173(82.8%) patients, Favipiravir in 59(28.2%), Remdisivir in 111(53.1%), Methylprednisolone in 87(41.6%), Dexamethasone in 93(44.5%), Antibiotics in 204(97.60%), Toccilizumab in 34(16.3%), plasma in 18(8.6%) and LMWH in 200(95.7%) patients. These were different treatment options provided the patients as per the then guideline of COVID 19 patients management in Bangladesh. Currently, no anti-viral agents have been proven to be an effective treatment for COVID-19. Remdesavir in hospitalized patients on oxygen was found to have reduced hospital stay but not mortality benefit and hence around 23% cases in this series received the drug. The study showed treatment of the patients with thromboprophylaxis, oxygen therapy (as needed), judicious use of steroid & antibiotics along with symptomatic management according to treatment guidelines was suffice.^15^

Regarding outcome of the COVID patients admitted, 85(92.4%) patients improved, 6(6.5%) died who were RT-PCR positive and 107(91.15%) improved, 9(7.7%) died who were probable cases. Death rate was little higher in our study then a study done before in Bangladesh which was 4.7%.^2^ Around two-third patients could be discharged in less than 10 days’ time, only few patients required longer duration of hospital stay (>30 days).

## Conclusion

According to this study, COVID-19 patients in Bangladesh have presenting symptoms like fever, cough, and berating complaints, nausea, vomiting, lethargy, and a higher temperature of >100°F. Male were more affected then female and middle age group are also more affected. Hematological findings like CRP was found to increase among all of our study patients. Besides, an increase in Serum ferritin, and D-Dimer along with erythrocytopenia and lymphocytopenia can be important supportive diagnostic criteria. Death rate is higher in our country and different treatment are applied as per national guideline which is changing with time.

## Data Availability

Yes

## Results

Table 1 showing most of the patients were male139(66.5%) and male to female ratio was 1.98:1.

**Table 1:** Gender distribution

|  | Frequency | Percent |
| --- | --- | --- |
| Male | 139 | 66.5 |
| Female | 70 | 33.5 |
| Total | 209 | 100.0 |

Table 2 showing age group distribution where patients aggregated in age group <20 years was 2(1.0%), 21-30 years were 9(4.3%), 31-40 years were 24(11.5%), 41-50 years 36(17.2%), 51-60 years were 54(25.8%) 61-70 years 57(27.3%) and >71 years 27(12.9%).

**Table 2:** Age group distributions

|  | Frequency | Percent |
| --- | --- | --- |
| <20 years | 2 | 1.0 |
| 21- 30 years | 9 | 4.3 |
| 31 - 40 years | 24 | 11.5 |
| 41- 50 years | 36 | 17.2 |
| 51- 60 years | 54 | 25.8 |
| 61- 70 years | 57 | 27.3 |
| >71 years | 27 | 12.9 |
| Total | 209 | 100.0 |

Table 3 showing 92(44%) patients were RT-PCR positive and 117(56%) were probable.

**Table 3:** Type of covid (n=209)

|  | Frequency | Percent |
| --- | --- | --- |
| RT-PCR confirmed covid | 92 | 44.0 |
| Probable covid | 117 | 56.0 |
| Total | 209 | 100.0 |

Table 4 showing clinical different features where fever was present in 195(93.3%) cases, cough in 180(86.1%), respiratory distress in 105(50.2%) anosmia in 123(58.8%), aguesea in 112(53.58%) and lethargy was present in 143(68.42%).

**Table 4:** Clinical features

|  | Frequency | Percent |
| --- | --- | --- |
| Fever | 195 | 93.3 |
| Cough | 180 | 86.1 |
| Res distress | 105 | 50.2 |
| Anosmia | 123 | 58.8 |
| Aguesea | 112 | 53.58 |
| Lethergy | 143 | 68.42 |

Table 5 showing Chest Xray findings where 43(20.6%) had normal findings, 73(34.9%) had bilateral patchy opacities, 20(9.6%) had unilateral opacities 65(31.1%) had consolidations, 6(2.9%) had ground glass opacities and 2(1.0%) had pleural effusion.

**Table 5:** CXR findings

|  | Frequency | Percent |
| --- | --- | --- |
| Normal | 43 | 20.6 |
| Bilateral patchy opacities | 73 | 34.9 |
| Unilateral patchy opacities | 20 | 9.6 |
| Consolidations | 67 | 32.1 |
| Ground glass opacities | 6 | 2.9 |
| Pleural effusion | 2 | 1.0 |
| Total | 209 | 100.0 |

Table 6 showing different treatment provided where supplemental O2 was given in 173(82.8%) patients, Favipiravir was given in 59(28.2%), Remdisivir was given in 111(53.1%), Methylprednisolone in 87(41.6%), Dexamethasone in 93(44.5%), Antibiotics was given in 204(97.60%), Toccilizumab in 34(16.3%), plasma in 18(8.6%) and LMWH in 200(95.7%) patients.

**Table 6:** Treatment provided

|  | Frequency | Percent |
| --- | --- | --- |
| O2 | 173 | 82.8 |
| Favipiravir | 59 | 28.2 |
| Remdisivir | 111 | 53.1 |
| Methylprednisolone | 87 | 41.6 |
| Dexamethasone | 93 | 44.5 |
| Antibiotics | 204 | 97.60 |
| Tocilizumab | 34 | 16.3 |
| Plasma | 18 | 8.6 |
| LMWH | 200 | 95.7 |

Table 7 showing different clinical and lab results

**Table 7:** Clinical and Lab data

|  | N | Minimum | Maximum | Mean | Std. Deviation |
| --- | --- | --- | --- | --- | --- |
| Pulse | 209 | 54 | 135 | 91.77 | 14.576 |
| Respiratory rate | 209 | 16 | 45 | 26.44 | 5.211 |
| SBP | 209 | 60 | 195 | 130.44 | 18.549 |
| DBP | 209 | 20 | 120 | 77.63 | 13.634 |
| O2 saturation on admission | 209 | 50 | 99 | 91.05 | 9.915 |
| Hemoglobin(gm/dl) | 200 | 7.00 | 15.50 | 12.1314 | 1.63307 |
| Total count | 194 | 1200 | 144000 | 12546.13 | 13889.207 |
| Neutrophil count | 196 | 36.0 | 93.0 | 75.162 | 10.4150 |
| Lymphocyte count | 195 | 3 | 80 | 20.43 | 11.179 |
| Platelet count | 188 | 30000 | 555000 | 268602.39 | 94021.263 |
| CRP | 209 | .90 | 304.70 | 62.4760 | 58.64221 |
| Ferritin | 209 | 1.42 | 3000.00 | 531.9388 | 474.99589 |
| LDH | 78 | 110.00 | 2700.00 | 420.7308 | 365.43199 |
| D-dimer | 209 | .05 | 67.00 | 1.6798 | 5.00399 |

Table 8 showing outcome of COVID patients admitted where 85(92.4%) patients improved, 6(6.5%) died who were RT-PCR positive and 107(91.15%) improved, 9(7.7%) died who were probable cases. Total death rate was 7.1%.

**Table 8:**
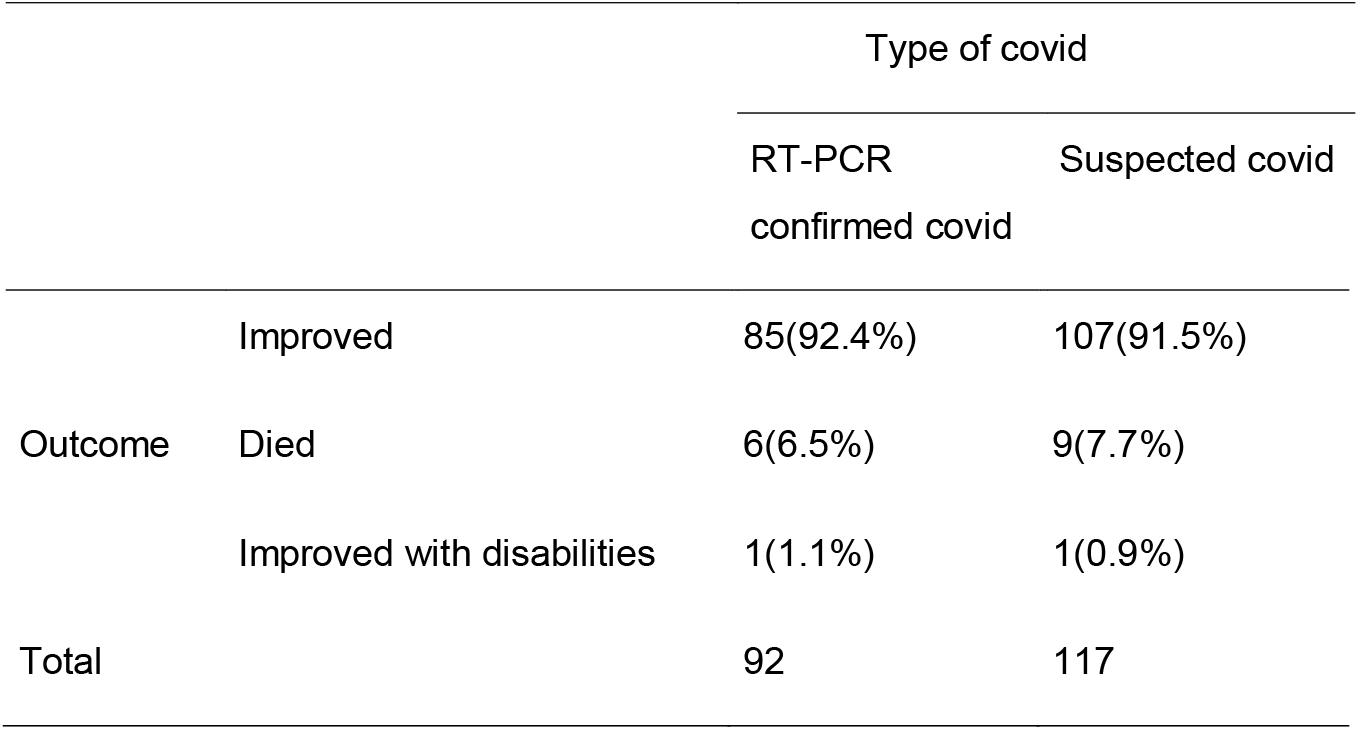
Relation of outcome with covid type

## References

1. Chen N, Zhou M, Dong X, Qu J, Gong F, Han Y. Epidemiological and clinical characteristics of 99 cases of 2019 novel coronavirus pneumonia in Wuhan, China: a descriptive study. Lancet. 2020; 15;395(10223):507–513.

2. Chowdhury Atmm, Karim MR, Mehedi HMH, Shahbaz M, Chowdhury MW, Dan G. Analysis of the primary presenting symptoms and hematological findings of COVID-19 patients in Bangladesh J Infect Dev Ctries 2021; 15(2):214–223

3. WHO, Bangladesh. Situation report coronavirus (COVID-19)-31. March 2021. https://www.who.int/bangladesh/emergencies/coronavirus-disease-(covid-19)-update/coronavirus-disease-(covid-2019)-bangladesh-situationreports

4. Guan W, Ni Z, Hu Y, et al. Clinical characteristics of coronavirus disease 2019 in China. N Engl J Med 2020. doi:10.1056/NEJMoa2002032

5. Mowla SGM, Azad KAK, Kabir A, Biswas S, Islam MT, Banik GC, Khan MH, Rohan KMI. Clinical Profile of 100 Confirmed COVID-19 Patients Admitted in Dhaka Medical College Hospital, Dhaka, Bangladesh J Bangladesh Coll Phys Surg 2020; 38: 29–36

6. Huang C. Wang Y, Li X et al. Clinical features of patients infected with 2019 novel coronavirus in 312 Wuhan, China, Lancet 2020.

7. Wang D, Hu B, Hu C et al. Clinical Characteristics of 138 Hospitalized patients with 2019 Novel Coronavirus infected Pneumonia in Wuhan, China. JAMA 2020

8. Guan WJ, Ni ZY, Hu Y, Liang WH, Ou CQ, He JX, et al. Clinical Characteristics of Coronavirus Disease 2019 in hina. N Engl J Med. 2020;382(18):1708-1720

9. Gupta N, Agrawal S, Ish P, Mishra S, Gaind R, Usha G, et al. Clinical and epidemiologic profile of the initial COVID-19 patients at a tertiary care centre in India. Monaldi Arch Chest Dis. 2020;90(1):193–6.

10. Hossain I, Khan MH, Rahman MS, Mullick AR, Aktaruzzaman MM. The Epidemiological Characteristics of an Outbreak of 2019 Novel Coronavirus Diseases (COVID-19) In Bangladesh: A Descriptive Study. JMSCR. 2020;08(04):544–551.

11. Chung M, Bernheim A, Mei X, et al. CT imaging features of 2019 novel coronavirus (2019-NCoV). Radiology. 2020;295:202–7.

12. Fang Y, Zhang H, Xie J, et al. Sensitivity of chest CT for COVID-19: comparison to RT-PCR. Radiology. 2020;19:200432

13. Wong HYF, Lam HYS, Fong AHT, et al. Frequency and distribution of chest radiographic findings in COVID-19 positive patients. Radiology. 2019;27:201160

14. Terpos E, Ntanasis-Stathopoulos I, Elalamy I, Kastritis E, Sergentaris TN, Politou M, Psaltopoulou T, Gerotziafas G,Dimopoulos MA. Hematological findings and complications of COVID-19. Am J Hematol 2020;95: 834–847.

15. Islam QT, Hossain HT, Fahim FR, Rashid MU. Clinico-Demograhic Profile, Treatment Outline And Clinical Outcome Of 236 Confirmed Hospitalized Covid-19 Patients: A Multi-Centered Descriptive Study In Dhaka, Bangladesh BJM 2020; 319 (2): 52–57

